# Disparities of Female Authorship in Colombia: A 5-year Cross-sectional Analysis

**DOI:** 10.1101/2023.09.22.23295823

**Authors:** Maria Alejandra Gutierrez-Torres, Silvana Ruiz, Karen Morales, Frans Serpa, Laura Rincon, Camila Gómez, Michelle M. Ahrens, Felipe Duran, Abul Ariza Manzano, Santiago Callegari

## Abstract

Accepted medical journal manuscripts serve as the primary currency of the scientific community. Over the past few decades, there has been a gradual increase in the number of women entering the medical field. However, women remain underrepresented as first and last authors in medical journals. This lack of representation makes it harder for them to reach leadership roles and advance in their academic careers. Therefore, this study aims to investigate the gender distribution among authors and explore disparities in authorship in relation to the type of publication (original research, case reports, and review articles) as well as the authors’ geographic locations. Our analysis included 6,088 articles from 54 research journals sourced from Colombia’s Ministry of Health official website. These articles were predominantly original research articles, although case reports and reviews were also present. The analysis specifically focused on published research articles, as they are extensively studied in existing literature. Until now, there has been no assessment of gender disparities in authorship within medical and surgical specialty journals in Colombia. We could evidence persistent gender disparity in primary authorship in Colombia, regardless of the timeframe, location, or field of study. This emphasizes the urgent need for enhanced support for female researchers and equitable resource allocation to rectify regional imbalances. Furthermore, our findings highlight the imperative need to address gender disparities in authorship in medical and surgical research articles in Colombia and other Latin American countries.

## Introduction

Accepted medical journal manuscripts serve as the gold standard in the scientific community. Since the introduction of evidence-based medicine in 1990, the importance of conducting research has grown significantly (1). Peer-reviewed journal articles play an important role in expanding careers in the health professions (2). Despite a recent increase in the number of women entering medicine and its subspecialties (3), there is a significant underrepresentation of women as first and last authors in medical journals (4). This underrepresentation limits their opportunities for leadership positions and academic advancement.

In Latin American countries, similar gender disparities in authorship and academia persist in the medical field. Tinoco et al. (5) discovered that female authors were less likely to hold first and last positions in Brazilian Cardiology journals. A subsequent investigation by Campos et al. (6) revealed that women were frequently underrepresented on editorial boards in Latin American surgical journals. Despite increased female representation in medical schools and medical specialties in Latin American countries such as Colombia over the last decade (7), gender disparities in authorship have not been properly investigated. These disparities are most likely the result of a combination of factors, such as societal norms, limited mentorship opportunities, and individual perceptions of work-life balance (8).

For instance, up to this point, no evaluation of gender disparities in authorship in medical and surgical specialty journals in Colombia has been conducted. As a result, the purpose of this study is to look into the gender distribution of authors who contribute to these journals. We also intend to investigate authorship disparities in relation to the type of publication (original research, case reports, and review articles) and the authors’ geographic locations. The findings of this study may contribute to encouraging a more in-depth discussion about gender inequalities in Colombian academia, as well as act as a catalyst to promote female representation and inclusivity in the medical fields of Latin American countries.

## Methods

### Criteria for Journal Selection

Journals in the biomedical sciences related to medicine that have been continuously published for two years or more were defined as inclusion criteria. In our analysis, we included both university-affiliated medical journals with a broad scope and specialty-specific medical journals. Journals in biological or healthcare sciences that did not primarily focus on medical sciences, such as veterinary science, biology, nursing, and dentistry, were excluded.

### Journal Search Methodology

To include all possible Colombian journals, Scielo, Directory of Open Access Journals, PubMed, and Publindex were screened. Given the limited number of Colombian journals in databases such as PubMed, we conducted a deeper search in order to ensure inclusivity, taking into account journals from various regions and specialties within Colombia. Publindex, a government-affiliated entity in charge of overseeing scientific journals in Colombia, was our primary source for identifying Colombian journals. We excluded journals that were not classified as medical sciences and any journals with titles indicating a focus other than medicine, such as biological sciences (e.g., veterinary, biology) or healthcare sciences (e.g., nursing, dentistry). A second author independently conducted the same protocol to ensure the rigor of our screening process, and only journals that received consensus from both authors were included in our analysis. Following that, we reviewed all of the selected journals to ensure that they focused exclusively on medicine. If any articles unrelated to medicine were identified during this review, the journal in question was excluded (**Figure 1**).

**Figure 1.**
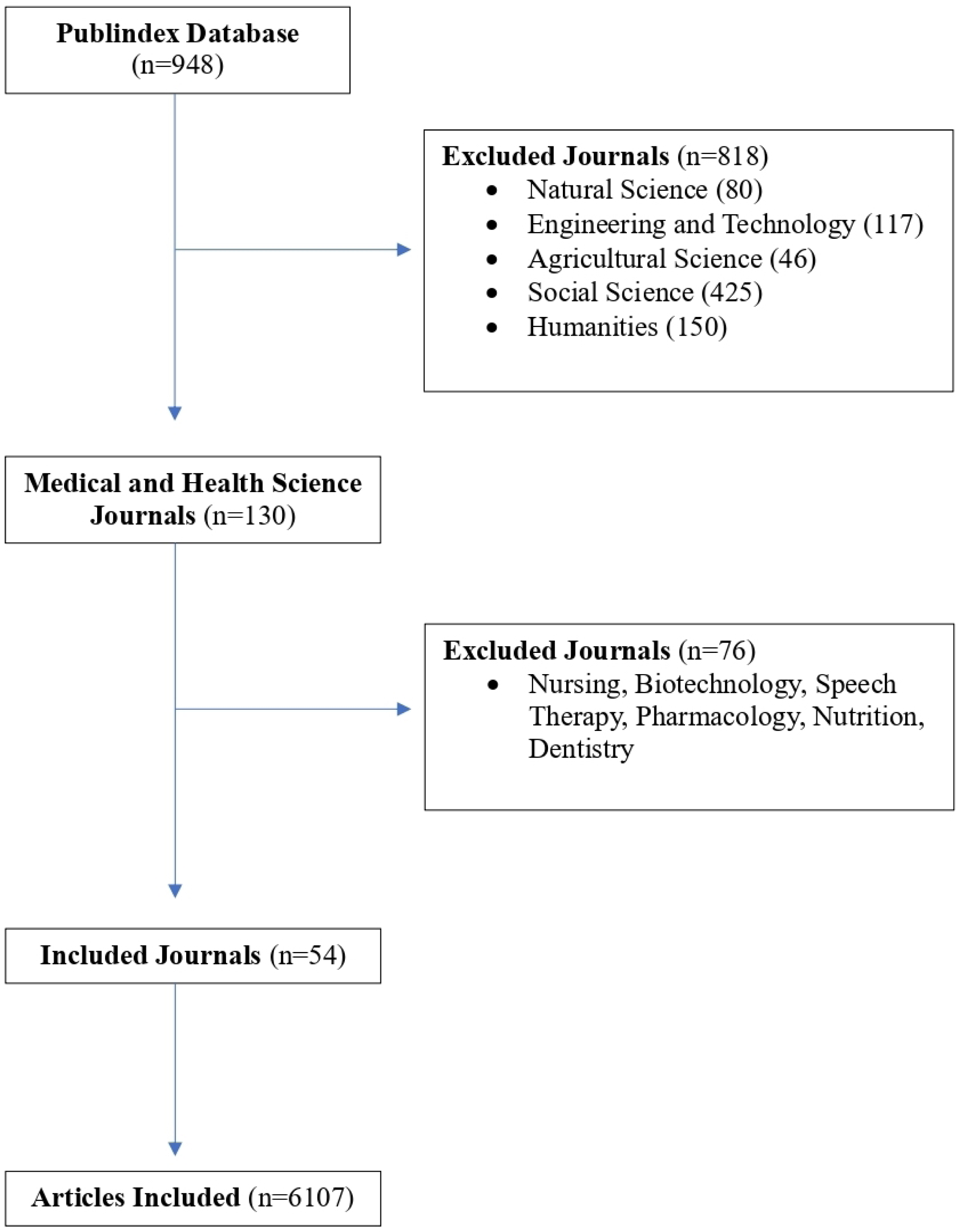
Flowchart of including and excluding scientific journals and articles for the analysis.

### Criteria for Article Selection

Articles were selected in accordance with the following inclusion criteria: 1) original research articles, review articles, and case studies; and 2) online public articles published in Colombian journals between 2018 and 2022. We excluded editorials, opinion pieces, special editions, and unaffiliated articles as they were not considered to be within the scope of our research. Furthermore, to avoid bias, we excluded journals that included articles from multiple specialties.

### Article abstractions

We conducted a cross-sectional study to determine the rates of authorship across multiple medical and surgical specialties. To accomplish this, we performed a bibliographic search that included all original articles published between January 1, 2018, and December 31, 2022, specifically in journals affiliated with Colombia’s leading medical associations.

We gathered information from the websites of each journal from January 2023 to May 2023, and we collected data from each journal’s website. We used the online Genderize webpage/application (https://genderize.io/) to determine the gender of the authors based on the names of the first and senior authors. Furthermore, we conducted Google searches for the authors’ names when required, cross-referencing the information with the official websites of their institutions. The software determines the gender of each name using a probabilistic method based on global published literature. The data was independently assessed by two or more researchers at each stage, and discrepancies were resolved by consensus. When there was doubt about the gender of the authors, we excluded the article from the statistical analysis.

Following the exclusion of articles, the researchers independently collected data from the remaining articles. The following data was extracted during this data collection process: article type, first author’s name, last author’s name, corresponding author’s name, and affiliations. After that, all collected data was consolidated and saved in a shared Microsoft Excel database. For our statistical analysis, we used Fisher’s exact test and calculated association measures using STATA 17 BE software. The level of significance we chose was 5%.

## Results

This study reviewed 54 journals obtained from the official website of Colombia’s Ministry of Health, specifically the journal regulation information available on publindex.com. The exclusion criteria were primarily focused on journals with broad coverage in biological or healthcare sciences (**Figure 1**). In total, 6,088 articles were included in our analysis. These articles were mostly original research articles, but there were also case studies and reviews, as shown in **Table 1**. The investigation was explicitly focused on research articles considering that they are extensively studied in the existing literature.

**Table 1.** Association of the first author being female when the last author is female.

| Authorship between first and last authors according to the type of article |  |  |  |  |
| --- | --- | --- | --- | --- |
| Type of article | Last Female Author | Last Male Author | First Female Author | First Male Author |
| Research article | 1322/3261 **(0.41)** | 1939/3261 **(0.59)** | 1504/3261 **(0.46)** | 1757/3261 **(0.54)** |
| Case Report | 569/1572 **(0.36)** | 1003/1572 **(0.64)** | 603/1572 **(0.38)** | 969/1572 **(0.62)** |
| Review Article | 535/1254 **(0.43)** | 719/1254 **(0.57)** | 573/1254 **(0.46)** | 681/1254 **(0.54)** |

### Female Authorship Over Time

In terms of author gender distribution, data collected indicates that female author participation has generally remained stable over time. The articles were organized chronologically based on their publication year, and some notable trends have been identified. Over the last five years, the number of female first authors has steadily increased. During this time, however, the number of female last authors has remained relatively constant. Similar patterns were observed when we analyzed reviews and case reports by publication timeline, as detailed in **Supplementary Tables 1 and 2**. Interestingly, female first authorships increased slightly in 2022 and 2019, but decreased in 2021. Similarly, last female authorships increased in 2021 but decreased in 2022 and 2019, indicating a possible inverse relationship, as illustrated in **Figure 2**.

**Figure 2.**
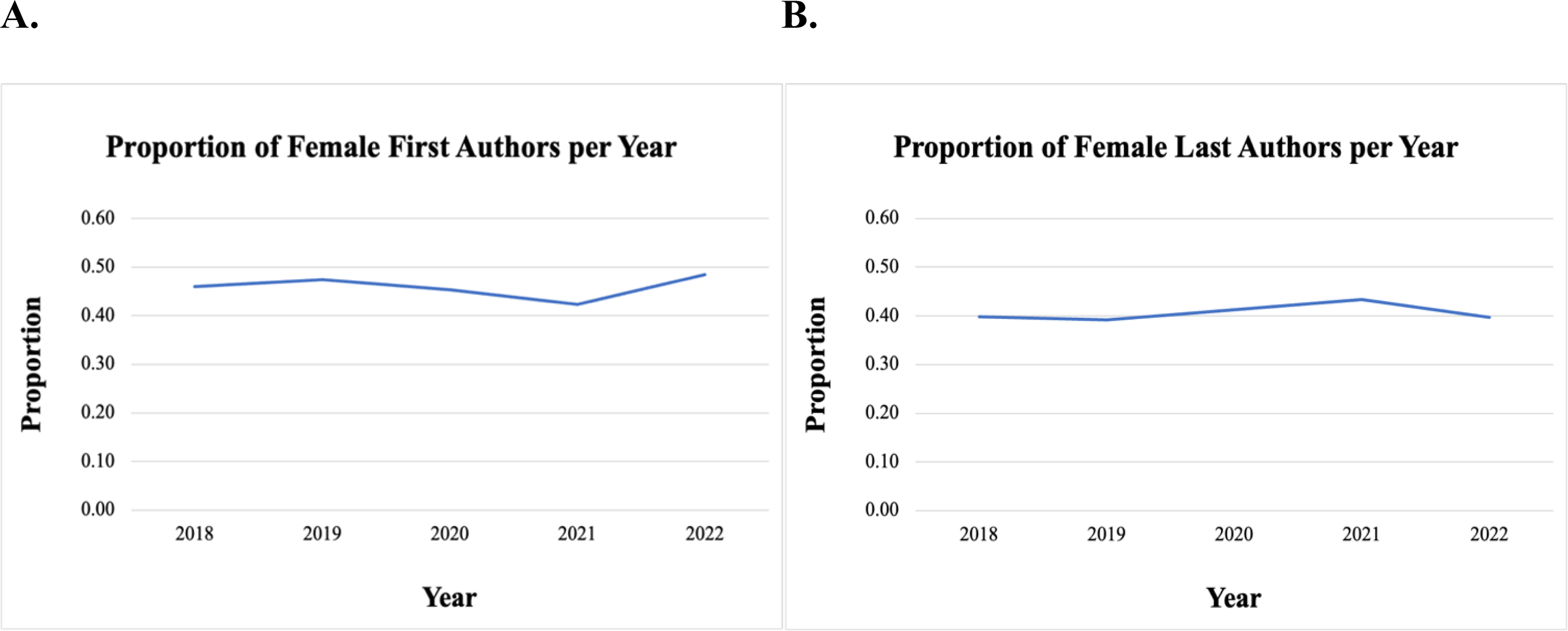
Proportion of female last and first authors per year.

### Colombian Female Authorship Rates Vary by Specialty

The journals were classified according to their primary focus, whether it was medicine, surgery, or a broader range of topics (miscellaneous). We also divided the journals into subspecialties, such as cardiology, nephrology, and plastic surgery, among others. When we studied research articles, we discovered significant differences in female author participation across journals, as shown in **Figure 3**. In general, surgical journals had fewer female authors than non-surgical journals, which is aligned with previous studies (4) (9). This trend was especially noticeable in orthopedic surgery, consistent with existing literature (10). As shown in **Figure 3**, the data additionally indicates that subspecialties such as neurosurgery, orthopedics, and general surgery had fewer female authors acting as first, last, and correspondence authors. However, there is an increasing number of more female authors in subspecialties such as nutrition, dermatology, radiology, pediatrics, and public health.

**Figure 3.**
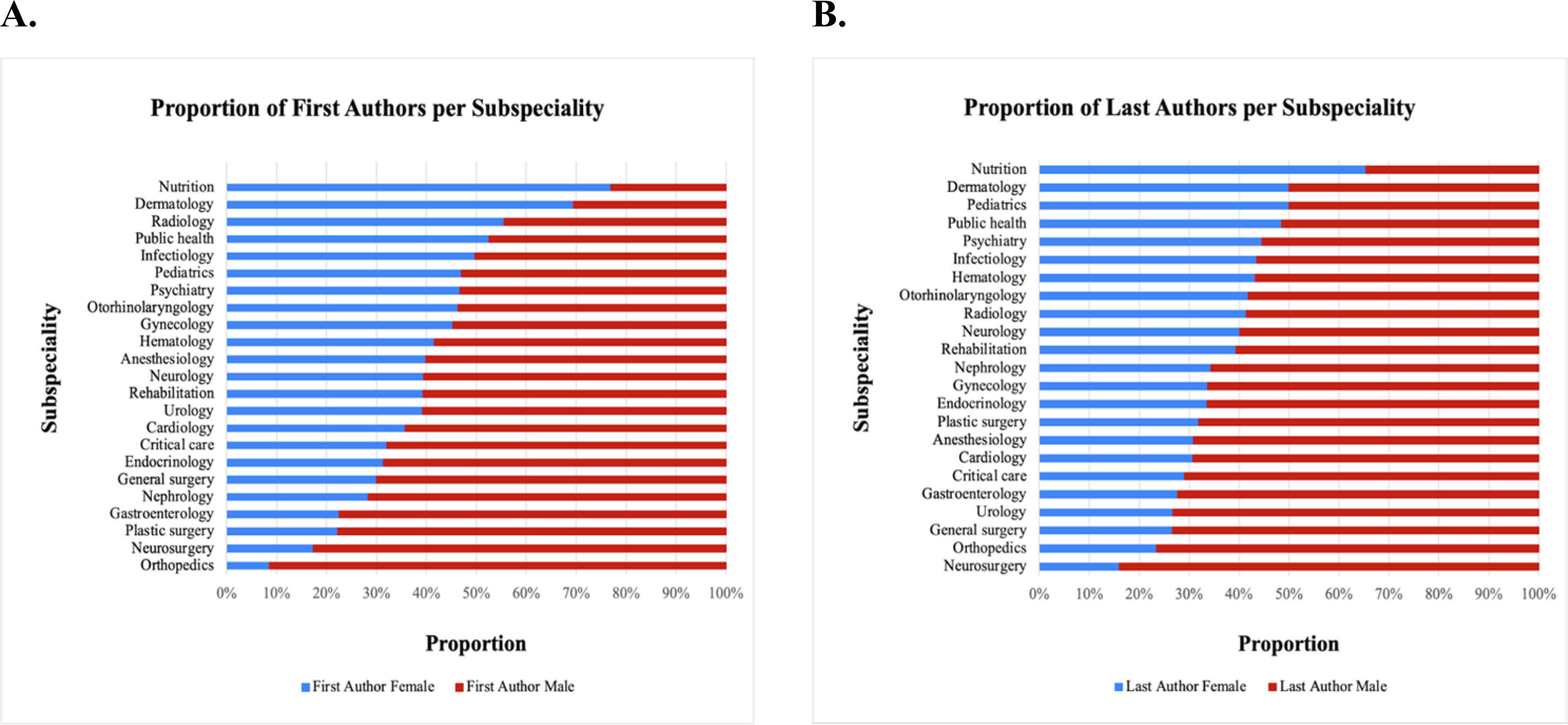
Proportion of female last and first authors per subspecialty.

### Colombian regions exhibit similar trends

Despite varying prevalence rates, the trend of lower female authorship remained consistent across regions. As shown in **Supplementary Figure 2**, the overwhelming majority of articles originated from Bogotá, Colombia’s capital city. We identified five primary regions that had a higher number of publication articles: Bogotá, Medellin, Cali, Bucaramanga, and Barranquilla, which include most of the country’s largest cities. Nonetheless, these were not the regions with higher female first and last female authors, revealing even more significant disparities across the regions of the nation. According to **Figure 4 A**, the proportion of female first authors was higher in regions such as Casanare, Santander, Caquetá, and Nariño.

**Figure 4.**
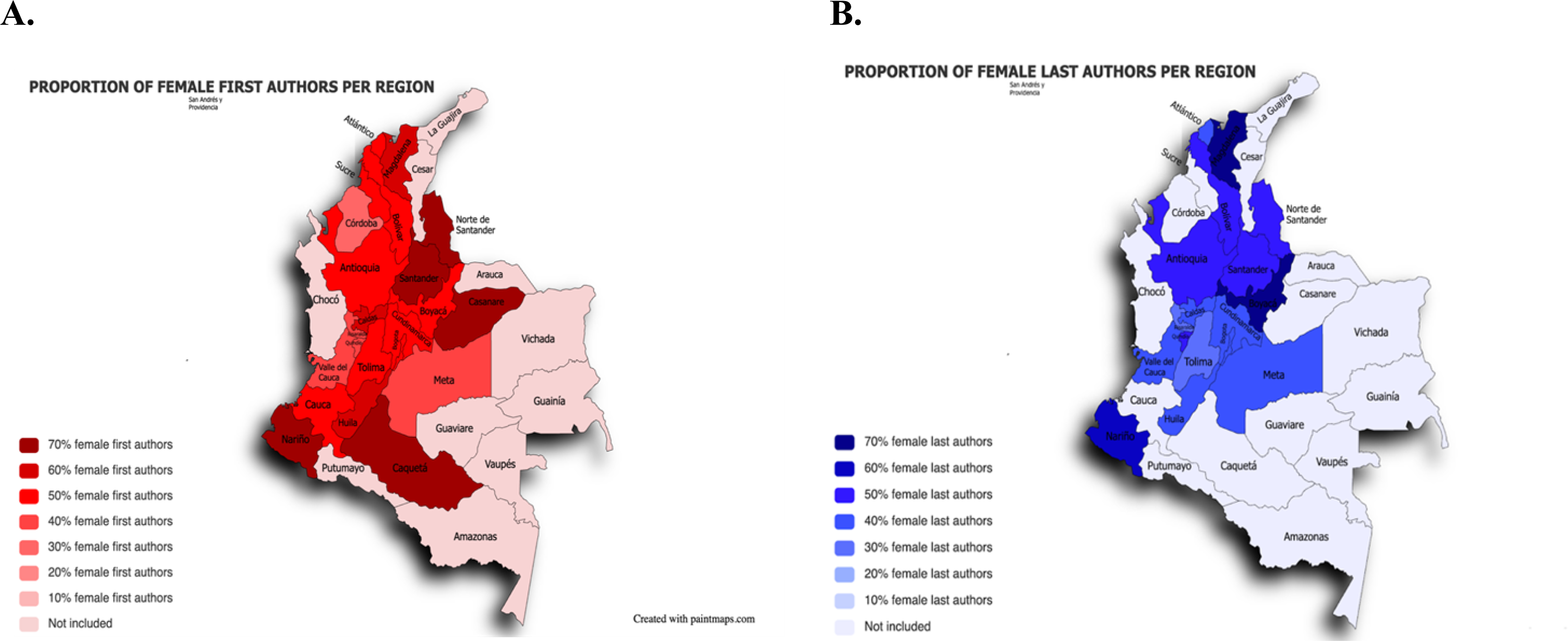
Proportion of female first (A) and last (B) authors per region.

In contrast, **Figure 4 B** illustrates the distribution of female last authors, showing a higher representation in Boyacá and Magdalena. Most regions, however, displayed similar patterns, highlighting a concentration of female-authored publications in the country’s central areas. It is also worth noting that the data revealed a greater proportion of regions with female first authors than female last authors.

### Association of Female First and Last Authors

We calculated association measures to gain a better understanding of the impact of female authors on first-author publications, as shown in **Table 2**. In line with previous research, our findings revealed a strong and statistically significant positive relationship between the gender of the first author and the gender of the last author in research articles. This pattern was evident not only in research papers but also in medical and surgical journals.

**Table 2.** Association of the first author being female when the last author is female.

| Association of the first author being female when the last author is female | OR | p value |
| --- | --- | --- |
| Research articles | 1.86 (IC95% 1.65-2.09) | p<0.0001 |
| Surgical articles | 0.82 (0.44-1.5) | p=0.518 |
| Medicine articles | 1.76 (1.46-2.11) | p<0.0001 |
| All articles included | 1.73 (IC95% 1.57-1.93) | p<0.0001 |

## Discussion

Gender disparities in authorship persist across various academic disciplines in Colombia, regardless of time or region (11). Female authorship in Colombia has remained stable over time, with surgical journals having fewer female authors than medical journals (11). Additionally, the presence of a female first author consistently aligns with the presence of a female last author, a pattern observed in both medical and surgical journals. Our study adds significant value to the existing literature by including a wide range of medical and surgical specialties and non-PubMed indexed journals, frequently excluded from traditional bibliometric studies. This incorporation is significant given that it expands the sources of knowledge and supports a wider spectrum of the academic and medical literature in Colombia.

Besides this, the profound methodology used in this study enabled us to investigate disparities across a large geographic area in Colombia, leading to a more comprehensive understanding of authorship trends in various geographic areas of the country. The distribution of authors in academic publications is highly dependent on the region and field of study. According to a study conducted in Colombia, only 12 of the 32 territorial departments demonstrated a significant volume of academic publications in the field of surgery with students as authors (12). This finding suggests that there may be disparities in research collaboration and authorship across Colombian regions. In our study, a similar pattern emerged, with only five regions in Colombia exhibiting a predominance of female authors. This highlights the need for more research into how authorship is distributed across Colombia’s various fields and regions. It is essential to explore the factors that may contribute to these disparities and gain a thorough understanding of the underlying causes.

According to our findings, in Colombia, female authorship has remained stable over time. While first authorship has increased slightly over the last five years, last authorship has remained consistent across all article types. Hart et al. (2013) examined the trend in female authorship across medical specialties in high-impact factor journals from 2008 to 2018. During the study period, there was a 3.6% increase in women as first authors and a 7.8% increase in women as last authors. They uncovered a higher presence of female first authors in articles where the last author was female, which is analogous to our results.

Nonetheless, in contrast to our findings, they identified a consistent increase in the proportion of female authors as last authors. However, this discrepancy could be related to what a subsequent study (14) identified, that the increase in female representation in surgical journals was less pronounced. Female last authors increased at roughly half the rate of female first authors in high-impact surgical journals. They discovered that the proportion of female last authors increased at a rate of 0.53% per year, while female first authorship increased at a faster rate of 0.97% per year. Female representation in specific journals, such as Liver Transplantation, European Journal of Cardiothoracic Surgery, and Journal of Vascular Surgery, was significantly lower in the last authorships. The findings of our study support these observations, highlighting a disparity in female representation in surgical journals. We did, however, notice a lower representation in the fields of neurosurgery, orthopedics, and general surgery.

Besides this, our findings reveal that females are significantly underrepresented in Colombian medical and surgical journals. The majority of articles with female first authors are concentrated in the country’s major urban centers, highlighting regional disparities in research opportunities and the need for investment and support in rural areas. This pattern is consistent with previous Latin American studies (5) (6), highlighting the continued underrepresentation of females as first and last authors in research articles. It has even been demonstrated that women’s representation is declining in Latin America, primarily in the medical sciences (11). Despite the slow but steady increase in female authorship worldwide, it is critical to recognize that low and middle-income countries face significant challenges (15). This emphasizes the importance of addressing the challenges that women face in this field and working toward gender parity. For instance, it is fundamental to enhance mentorship to promote women’s professional development in academia across the region (11). Also, considering what was evidenced in Colombia, future research should investigate whether similar gender disparities in authorship exist in other Latin American countries, as well as the factors that contribute to these disparities. This could be the first step to decreasing gender disparities in medical authorships.

Despite the fact that the gender disparity in authorship has been widely published, the majority of the studies are from journals in the United States of America. Nonetheless, a search of statistics in Latin America revealed minimal available literature on this subject, with only two studies identified. Graner et al. conducted one in Brazil and discovered that there is an underrepresentation of women in surgical Brazilian journals, with an increasing number of women as first authors and fewer as last authors. Furthermore, they demonstrated that orthopedics is one of the subspecialties with the greatest underrepresentation (17). The other study in pediatrics by Dominguez et al. with journals from Argentina and Chile found that, while women’s authorship has increased over time, they still need to be represented and frequently appear as the last authors (18). Similarly, post-pandemic global research in scientific publications, including the medical field, revealed that Latin America had the lowest percentage of female authorship, with 16%, when compared to Asia, Africa, Europe, and Oceania (19).

Gender biases, cultural norms, and unequal access to education and career advancement may all contribute to the underrepresentation of female authors in Latin America. Eliminating these barriers and implementing gender-equitable policies is crucial for fostering an inclusive research environment (16). This effort could include programs like female mentorship, promoting work-life balance, and enforcing policies that ensure equal career opportunities. In addition, concerted efforts must be made to empower and support researchers in underrepresented regions by supplying them with the necessary resources and infrastructure for meaningful research engagement. This assistance could originate in the form of establishing research centers or partnerships in these areas, as well as facilitating networking opportunities to improve knowledge exchange.

### Study limitations

We acknowledge the inherent complexity of our research problem and the limitations of our investigation. First and foremost, because we used a manual article selection process, it is critical to recognize the possibility of human error. However, the large number of articles included in our study increases our confidence in the validity of our findings. Second, we needed to exclude specific journals that were outside the scope of medicine. However, deliberate efforts were made to ensure representation from key regions in Colombia, thereby increasing the depth of our research. Although our temporal analyses did not reveal any significant deviations, we must consider the potential impact of the COVID-19 pandemic on evolving trends. While our study was limited to Colombia, it is critical to recognize the need for similar initiatives in fields beyond of the boundaries of our research. Finally, we recognize that some of the journals we investigated may have needed more coverage of specific subspecialties, which could introduce bias. Nonetheless, as previously demonstrated, including subspecialty-focused journals provides a more accurate picture of female authorship trends. This aspect distinguishes our approach, especially when compared to academic publications that are primarily focused on educational contexts rather than clinical settings.

## Conclusion

In conclusion, regardless of timeframe, location, or field of study, our study reveals a persistent gender disparity in primary authorship in Colombia. This highlights the critical need for increased support for female researchers as well as equitable resource allocation to correct regional imbalances. Our comprehensive analysis offers practical recommendations and significantly contributes to ongoing efforts to create a more equitable research landscape. Additionally, our findings highlight the critical need to address gender disparities in medical and surgical research article authorship in Colombia and other Latin American countries. Despite its limitations, our study enriches the existing body of written work. It promotes collaborative efforts toward developing an inclusive research environment that recognizes and values the contributions of all researchers, regardless of gender or ethnicity. Collective action and sustained research efforts can lead to progress.

## Acknowledgments

Dra. Martha Gulati for her guidance and support on this project.

## Funding

The author(s) received no specific funding for this work.

## Competing Interests

The authors have declared that no competing interests exist.

## Data availability

All the data is publicly available without restrictions upon reasonable request to the authors.

## Supplementary figures

**Supplementary Figure 1.**
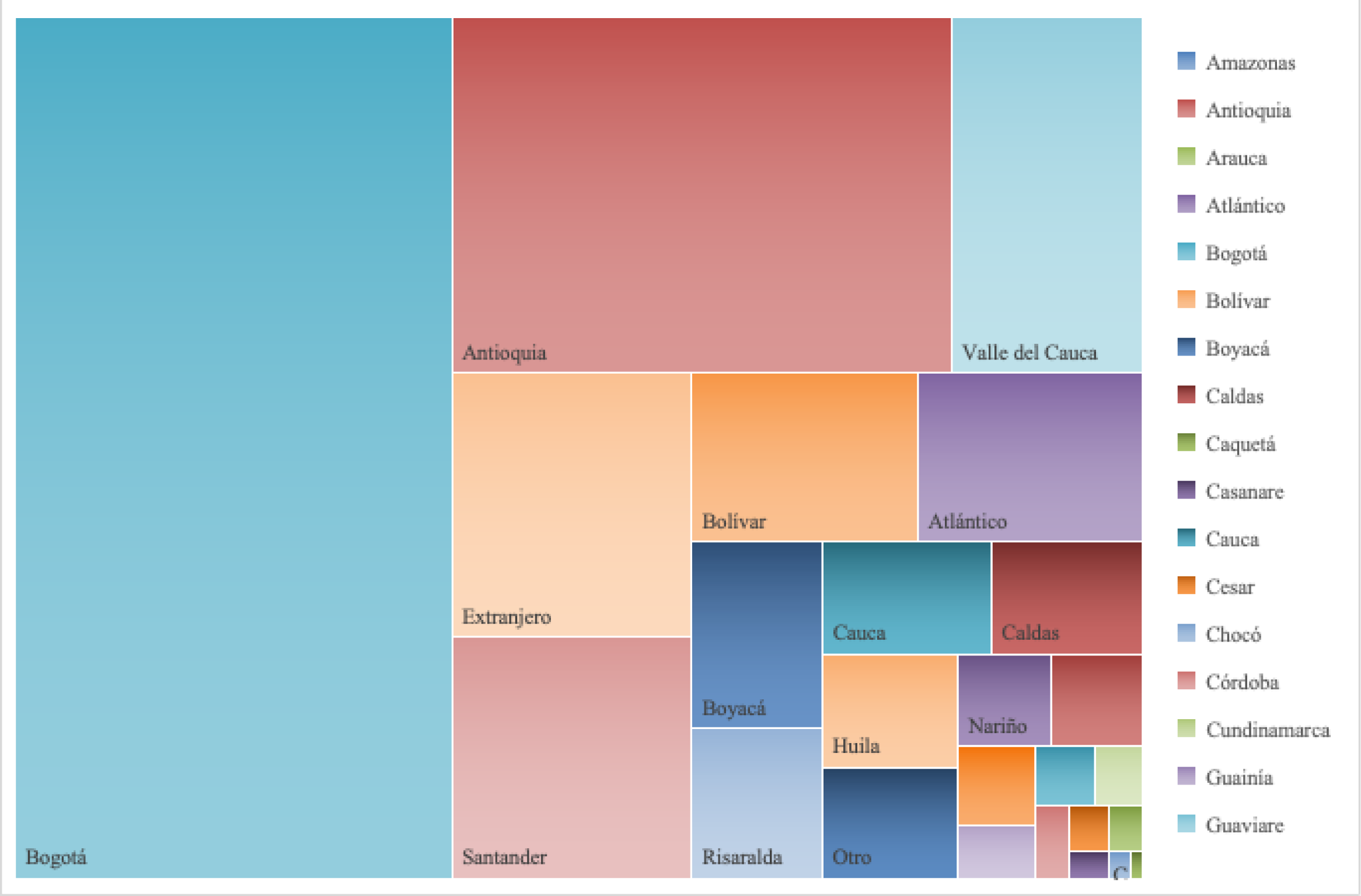
Number of articles per region.

**Supplementary Table 1.**
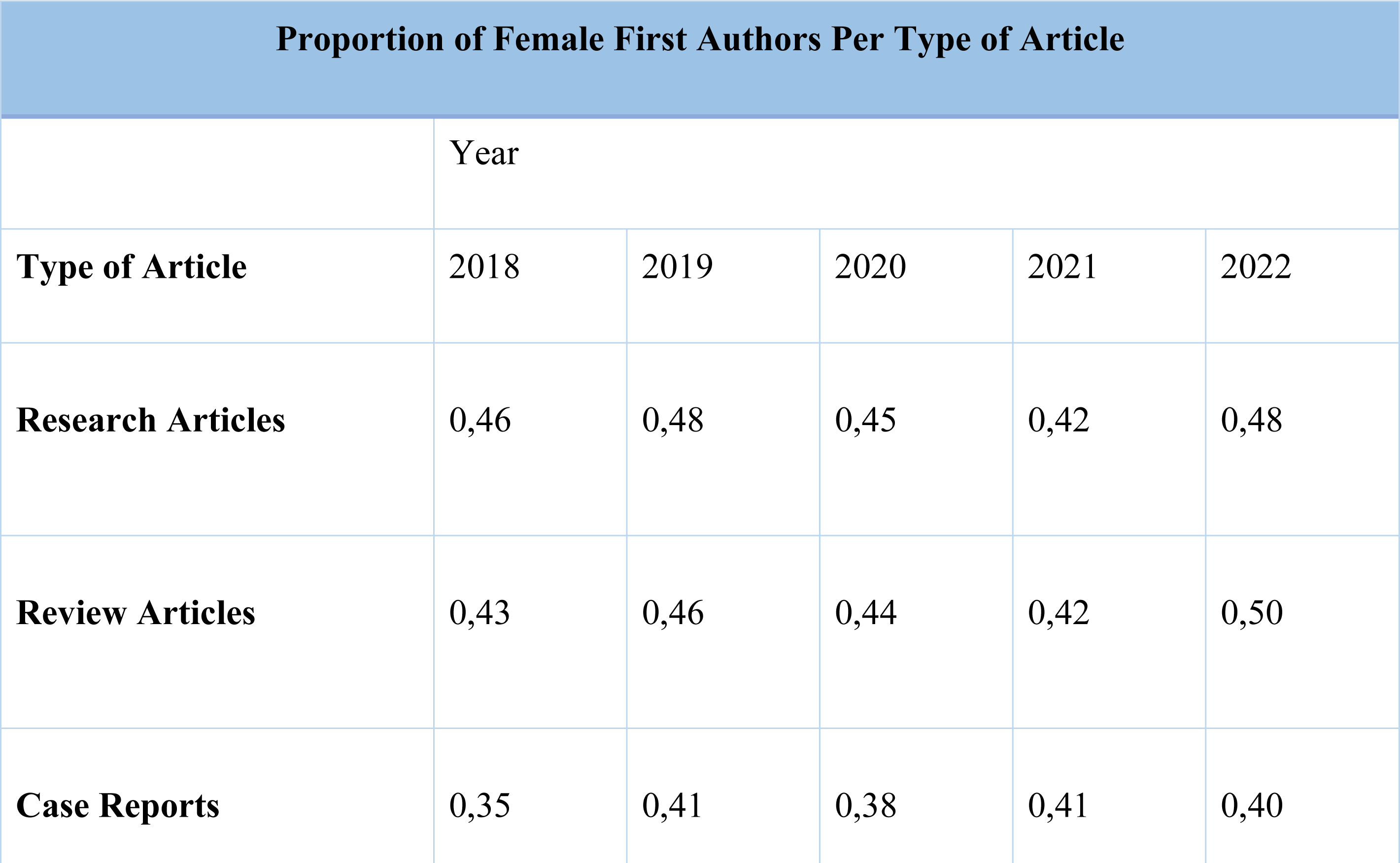
Female first authorship per type of article.

**Supplementary Table 2.** Female last authorship per type of article.

| Proportion of Female Last Authors Per Year |  |  |  |  |  |
| --- | --- | --- | --- | --- | --- |
|  | Year |  |  |  |  |
| Type of Article | 2018 | 2019 | 2020 | 2021 | 2022 |
| Research Articles | 0,40 | 0,39 | 0,41 | 0,43 | 0,40 |
| Review Articles | 0,41 | 0,44 | 0,45 | 0,42 | 0,42 |
| Case Reports | 0,34 | 0,35 | 0,38 | 0,41 | 0,33 |

**Supplementary Table 3.** Female authorship per subspeciality.

| Proportion of Female Authors Per Subspecialty |  |  |  |  |  |  |
| --- | --- | --- | --- | --- | --- | --- |
| Subspecialty | First author |  | Last author |  | Correspondence author |  |
|  | Female | Male | Female | Male | Female | Male |
| Anesthesiology | 0,3984 | 0,6016 | 0,3089 | 0,6911 | 0,4228 | 0,5772 |
| Neurosurgery | 0,1733 | 0,8267 | 0,16 | 0,84 | 0,1333 | 0,8667 |
| Orthopedics | 0,0851 | 0,9149 | 0,234 | 0,766 | 0,2411 | 0,7589 |
| Gynecology | 0,4535 | 0,5465 | 0,3372 | 0,6628 | 0,4302 | 0,5698 |
| Public health | 0,5256 | 0,4744 | 0,4846 | 0,5154 | *No data* |  |
| Nutrition | 0,7692 | 0,2308 | 0,6538 | 0,3462 | 0,7308 | 0,2692 |
| Nephrology | 0,2828 | 0,7172 | 0,3434 | 0,6566 | 0,2727 | 0,7273 |
| Otorhinolaryngology | 0,4636 | 0,5364 | 0,4182 | 0,5818 | 0,4862 | 0,5138 |
| General surgery | 0,3 | 0,7 | 0,2667 | 0,7333 | 0,2667 | 0,7333 |
| Critical care | 0,3212 | 0,6788 | 0,2909 | 0,7091 | 0,3232 | 0,6768 |
| Gastroenterology | 0,2248 | 0,7752 | 0,2769 | 0,7231 | 0,2117 | 0,7524 |
| Pediatrics | 0,47 | 0,53 | 0,5 | 0,5 | 0,51 | 0,49 |
| Plastic surgery | 0,2222 | 0,7778 | 0,3194 | 0,6806 | *No data* |  |
| Dermatology | 0,6939 | 0,3061 | 0,5 | 0,5 | 0,7143 | 0,2857 |
| Urology | 0,392 | 0,608 | 0,267 | 0,733 | 0,4057 | 0,5943 |
| Hematology | 0,416 | 0,584 | 0,432 | 0,568 | 0,432 | 0,568 |
| Neurology | 0,3942 | 0,6058 | 0,4015 | 0,5985 | 0,3723 | 0,6277 |
| Endocrinology | 0,3139 | 0,6861 | 0,3358 | 0,6642 | 0,3594 | 0,6406 |
| Cardiology | 0,3576 | 0,6424 | 0,3079 | 0,6921 | 0,298 | 0,702 |
| Radiology | 0,5556 | 0,4444 | 0,4141 | 0,5859 | 0,5567 | 0,4433 |
| Psychiatry | 0,4674 | 0,5326 | 0,4457 | 0,5543 | 0,4457 | 0,5543 |
| Infectiology | 0,4969 | 0,5031 | 0,4348 | 0,5652 | 0,4969 | 0,5031 |
| Physical medicine and<br>rehabilitation | 0,3934 | 0,6066 | 0,3934 | 0,6066 | 0,4262 | 0,5738 |

**Supplementary Table 4.** Female authorship per journal.

| Proportion of Female Authors Per Journal |  |  |  |  |  |  |
| --- | --- | --- | --- | --- | --- | --- |
| Journal | First author |  | Last author |  | Correspondence author |  |
|  | Female | Male | Female | Male | Female | Male |
| Colombian Journal of Anesthesiology | 0,3984 | 0,6016 | 0,3089 | 0,6911 | 0,4228 | 0,5772 |
| Neurociencias Journal | 0,1733 | 0,8267 | 0,16 | 0,84 | 0,1333 | 0,8667 |
| Revista Colombiana de Ortopedia y Traumatologia | 0,0851 | 0,9149 | 0,234 | 0,766 | 0,2411 | 0,7589 |
| Revista de Nutrición Clínica y Metabolismo | 0,5 | 0,5 | 0,6136 | 0,3864 | 0,4545 | 0,5455 |
| Medicina U.P.B. | 0,3889 | 0,6111 | 0,4667 | 0,5333 | 0,4333 | 0,5667 |
| Revista de la Facultad de Ciencias de la Salud | 0,4878 | 0,5122 | 0,5366 | 0,4634 | 0,6098 | 0,3902 |
| Revista Colombiana de Obstetricia y Ginecología | 0,4535 | 0,5465 | 0,3372 | 0,6628 | 0,4302 | 0,5698 |
| Salud Uninorte | 0,5031 | 0,4969 | 0,4417 | 0,5583 | 0,454 | 0,546 |
| Medicina y laboratorio | 0,6 | 0,4 | 0,4667 | 0,5333 | 0,6 | 0,4 |
| Biomédica: revista del Instituto Nacional de Salud | 0,5364 | 0,4636 | 0,4373 | 0,5627 | 0,4927 | 0,5073 |
| Colombia Médica | 0,3661 | 0,6339 | 0,2411 | 0,7589 | 0,3036 | 0,6964 |
| Revista Facultad Nacional de Salud Pública | 0,49 | 0,51 | 0,49 | 0,51 | *No data* |  |
| Perspectivas en Nutrición Humana | 0,7692 | 0,2308 | 0,6538 | 0,3462 | 0,7308 | 0,2692 |
| Universidad y Salud | 0,5935 | 0,4065 | 0,5528 | 0,4472 | 0,6098 | 0,3902 |
| Revista Colombiana de Nefrología | 0,2828 | 0,7172 | 0,3434 | 0,6566 | 0,2727 | 0,7273 |
| Revista Ciencias de la Salud | 0,5143 | 0,4857 | 0,3857 | 0,6143 | 0,4571 | 0,5429 |
| Acta De Otorrinolaringología & Cirugía De Cabeza Y Cuello | 0,4636 | 0,5364 | 0,4182 | 0,5818 | 0,4862 | 0,5138 |
| Revista Ciencias Biomédicas | 0,5072 | 0,4928 | 0,3913 | 0,6087 | 0,5797 | 0,4203 |
| Med UNAB | 0,5213 | 0,4787 | 0,5957 | 0,4043 | 0,4894 | 0,5106 |
| Acta médica colombiana | 0,3987 | 0,6013 | 0,3101 | 0,6899 | 0,3481 | 0,6519 |
| Revista Colombiana de Cirugía | 0,3 | 0,7 | 0,2667 | 0,7333 | 0,2667 | 0,7333 |
| Revista Ces Salud Pública | 0,2677 | 0,7323 | 0,2576 | 0,7424 | 0,2755 | 0,7245 |
| Biociencias | 0,5068 | 0,4932 | 0,3836 | 0,6164 | 0,4658 | 0,5342 |
| Revista de salud publica | 0,544 | 0,456 | 0,4819 | 0,5181 | *No data* |  |
| Ces Medicina | 0,4809 | 0,5191 | 0,4504 | 0,5496 | 0,4733 | 0,5267 |
| Revista Investigación en Salud | 0,8125 | 0,1875 | 0,6875 | 0,3125 | 0,7763 | 0,2237 |
| Acta Colombiana de Cuidado Intensivo | 0,3212 | 0,6788 | 0,2909 | 0,7091 | 0,3232 | 0,6768 |
| Revista Colombiana de Gastroenterología | 0,2248 | 0,7752 | 0,2769 | 0,7231 | 0,2117 | 0,7524 |
| Pediatría | 0,47 | 0,53 | 0,5 | 0,5 | 0,51 | 0,49 |
| Revista Colombiana de Cirugía Plástica y Reconstructiva | 0,2222 | 0,7778 | 0,3194 | 0,6806 | *No data* |  |
| Revista De La Asociación Colombiana De Dermatología Y Cirugía Dermatológica | 0,6939 | 0,3061 | 0,5 | 0,5 | 0,7143 | 0,2857 |
| Urología Colombiana | 0,392 | 0,608 | 0,267 | 0,733 | 0,4057 | 0,5943 |
| Revista Colombiana de Cancerología | 0,416 | 0,584 | 0,432 | 0,568 | 0,432 | 0,568 |
| Acta Neurológica Colombiana | 0,3942 | 0,6058 | 0,4015 | 0,5985 | 0,3723 | 0,6277 |
| Archivos de medicina | 0,3187 | 0,6813 | 0,2637 | 0,7363 | 0,3297 | 0,6703 |
| Revista de la Universidad Industrial de Santander. Salud | 0,6 | 0,4 | 0,5313 | 0,4688 | 0,5633 | 0,4367 |
| Revista Colombiana De Endocrinologia Diabetes Metabolismo | 0,3139 | 0,6861 | 0,3358 | 0,6642 | 0,3594 | 0,6406 |
| Revista Colombiana de Cardiología | 0,3576 | 0,6424 | 0,3079 | 0,6921 | 0,298 | 0,702 |
| Revista Universitas Medica | 0,5623 | 0,4377 | 0,4226 | 0,5774 | 0,5709 | 0,4291 |
| Revista Colombiana de Medicina Fisica y Rehabilitacion | 0,5962 | 0,4038 | 0,4808 | 0,5192 | 0,6346 | 0,3654 |
| Repertorio de Medicina y Cirugía | 0,3256 | 0,6744 | 0,3566 | 0,6434 | 0,3488 | 0,6512 |
| Duazary | 0,6226 | 0,3774 | 0,6132 | 0,3868 | 0,6132 | 0,3868 |
| Revista de la Facultad de Medicina | 0,4649 | 0,5351 | 0,4912 | 0,5088 | 0,4123 | 0,5877 |
| Revista Colombiana de Radiología | 0,5556 | 0,4444 | 0,4141 | 0,5859 | 0,5567 | 0,4433 |
| Revista Colombiana de Psiquiatría | 0,4674 | 0,5326 | 0,4457 | 0,5543 | 0,4457 | 0,5543 |
| Revista médica Sanitas | 1 | 0 | 1 | 0 | 0,3939 | 0,6061 |
| Revista Colombiana de Oftalmología | 0,55 | 0,45 | 0,4 | 0,6 | 0,7 | 0,3 |
| Revista Digital Actividad Física Y Deporte | 0,2429 | 0,7571 | 0,3286 | 0,6714 | 0,2714 | 0,7286 |
| Ciencia e Innovación en Salud | 0,5263 | 0,4737 | 0,4868 | 0,5132 | 0,4474 | 0,5526 |
| Medicina UIS | 0,4483 | 0,5517 | 0,3966 | 0,6034 | 0,4483 | 0,5517 |
| Revista Médica de Risaralda | 0,4902 | 0,5098 | 0,4314 | 0,5686 | 0,4706 | 0,5294 |
| Revista Navarra Médica | 0,7 | 0,3 | 0,65 | 0,35 | 0,7 | 0,3 |
| Infectio | 0,4969 | 0,5031 | 0,4348 | 0,5652 | 0,4969 | 0,5031 |

**Supplementary Table 5.** Female first authorship per region.

| Proportion Of Female First Authors Per Region |  |  |  |  |  |
| --- | --- | --- | --- | --- | --- |
| Region | Cities included in each region | Number of research articles per city | Number of research articles with first female author | Proportion of female first authors per city | Proportion per region |
| **Bogotá** | Bogotá | 1238 | 565 | 0,4564 |  |
|  | Chía | 8 | 5 | 0,625 |  |
|  | Soacha | 1 | 0 | 0 |  |
|  |  |  |  |  | **0,4571** |
| **Antioquia** | Medellín | 593 | 279 | 0,4705 |  |
|  | Sabaneta | 1 | 0 | 0 |  |
|  |  |  |  |  | **0,4697** |
| **Valle del Cauca** | Cali | 270 | 101 | 0,3741 |  |
|  | Palmira | 1 | 1 | 1 |  |
|  |  |  |  |  | **0,3764** |
| **Santander** | Bucaramanga | 174 | 106 | 0,6092 |  |
|  | Floridablanca | 3 | 1 | 0,3333 |  |
|  | Pamplona | 3 | 2 | 0,6667 |  |
|  |  |  |  |  | **0,6056** |
| **Bolívar** | Cartagena | 104 | 44 | 0,4231 |  |
|  |  |  |  |  | **0,4231** |
| **Atlántico** | Barranquilla | 134 | 62 | 0,4627 |  |
|  |  |  |  |  | **0,4627** |
| **Boyacá** | Tunja | 95 | 48 | 0,5053 |  |
|  | Sogamoso | 1 | 0 | 0 |  |
|  |  |  |  |  | **0,5** |
| **Cauca** | Popayán | 55 | 23 | 0,4182 |  |
|  |  |  |  |  | **0,4182** |
| **Risaralda** | Pereira | 65 | 22 | 0,3385 |  |
|  |  |  |  |  | **0,3385** |
| **Huila** | Neiva | 49 | 25 | 0,5102 |  |
|  |  |  |  |  | **0,5102** |
| **Magdalena** | Santa Marta | 38 | 20 | 0,5263 |  |
|  |  |  |  |  | **0,5263** |
| **Caldas** | Manizales | 33 | 17 | 0,5152 |  |
|  |  |  |  |  | **0,5152** |
| **Nariño** | Pasto | 30 | 21 | 0,7 |  |
|  |  |  |  |  | **0,7** |
| **Quindío** | Armenia | 31 | 10 | 0,3226 |  |
|  |  |  |  |  | **0,3226** |
| **Tolima** | Ibagué | 18 | 9 | 0,5 |  |
|  |  |  |  |  | **0,5** |
| **Norte de Santander** | Cúcuta | 11 | 7 | 0,6364 |  |
|  |  |  |  |  | **0,6364** |
| **Sucre** | Sincelejo | 10 | 7 | 0,7000 |  |
|  |  |  |  |  | **0,7** |
| **Meta** | Villavicencio | 8 | 3 | 0,3750 |  |
|  |  |  |  |  | **0,3750** |
| **Córdoba** | Montería | 7 | 2 | 0,3 |  |
|  |  |  |  |  | **0,30** |
| **Casanare** | Yopal | 2 | 1 | 0,5 |  |
|  |  |  |  |  | **0,5** |
| **Other** |  | 50 | 24 | 0,48 | **0,48** |
| **Foreign** |  | 169 | 60 | 0,355 | **0,355** |

**Supplementary Table 6.** Female last authorship per region.

| Proportion Of Female Last Authors Per Region |  |  |  |  |  |
| --- | --- | --- | --- | --- | --- |
| Region | Cities included in each region | Number of research articles per city | Number of research articles with female last author | Proportion of female last authors per city | Proportion per region |
| **Bogotá** | Bogotá | 1252 | 484 | 0,3866 |  |
|  | Chía | 9 | 5 | 0,5556 |  |
|  | Soacha | 1 | 0 | 0 |  |
|  |  |  |  |  | **0,3875** |
| **Antioquia** | Medellín | 591 | 242 | 0,4095 |  |
|  | Sabaneta | 1 | 0 | 0 |  |
|  |  |  |  |  | **0,4088** |
| **Valle del Cauca** | Cali | 258 | 87 | 0,3372 |  |
|  | Palmira | 1 | 0 | 0 |  |
|  |  |  |  |  | **0,3359** |
| **Santander** | Bucaramanga | 172 | 80 | 0,4651 |  |
|  | Floridablanca | 2 | 1 | 0,5 |  |
|  | Pamplona | 3 | 1 | 0,3333 |  |
|  |  |  |  |  | **0,4633** |
| **Bolívar** | Cartagena | 108 | 52 | 0,4815 |  |
|  |  |  |  |  | **0,4815** |
| **Atlántico** | Barranquilla | 120 | 42 | 0,35 |  |
|  |  |  |  |  | **0,35** |
| **Boyacá** | Tunja | 89 | 57 | 0,6404 |  |
|  | Sogamoso | 1 | 0 | 0 |  |
|  |  |  |  |  | **0,6333** |
| **Cauca** | Popayán | 54 | 28 | 0,5185 |  |
|  |  |  |  |  | **0,5185** |
| **Risaralda** | Pereira | 65 | 19 | 0,2923 |  |
|  |  |  |  |  | **0,2923** |
| **Huila** | Neiva | 48 | 19 | 0,3958 |  |
|  |  |  |  |  | **0,3958** |
| **Magdalena** | Santa Marta | 40 | 26 | 0,65 |  |
|  |  |  |  |  | **0,65** |
| **Caldas** | Manizales | 32 | 11 | 0,3438 |  |
|  |  |  |  |  | **0,3438** |
| **Nariño** | Pasto | 25 | 13 | 0,52 |  |
|  |  |  |  |  | **0,52** |
| **Quindío** | Armenia | 32 | 14 | 0,4375 |  |
|  |  |  |  |  | **0,4375** |
| **Tolima** | Ibagué | 14 | 3 | 0,2143 |  |
|  |  |  |  |  | **0,2143** |
| **Norte de Santander** | Cúcuta | 13 | 6 | 0,4615 |  |
|  |  |  |  |  | **0,4615** |
| **Sucre** | Sincelejo | 9 | 3 | 0,333 |  |
|  |  |  |  |  | **0,333** |
| **Córdoba** |  | 4 | 2 | 0,5 |  |
|  |  |  |  |  | **0,5** |
| **Meta** | Villavicencio | 5 | 1 | 0,2 |  |
|  |  |  |  |  | **0,2** |
| **Other** |  | 40 | 11 | 0,275 | **0,275** |
| **Foreign** |  | 199 | 70 | 0,3518 | **0,3518** |

## Notes

### Competing Interest Statement

The authors have declared no competing interest.

